# Planned care accounts for one in three recurrent hospitalizations in alcohol-related cirrhosis

**DOI:** 10.64898/2026.10.07.26365017

**Authors:** Krystian Mirowski, Aleksandra Zmuda, Hanna Herashchenko, Dorota Łukaszewska, Barbara Balicka-Slusarczyk, Piotr Hydzik, Michal Kukla

## Abstract

Hospital readmission is used in cirrhosis research as if a return to hospital meant the disease had worsened: it is a trial outcome, a quality indicator and a prediction target. We asked what those readmissions were. Sixty-one adults with alcohol-related cirrhosis, selected because they had been hospitalized more than once, contributed all 326 later admissions to one department in Krakow, Poland, between 2013 and 2026. A clinician read each full record and classified it for acute decompensation and for admission mode, blinded to the structured decompensation flags. Fewer than two in five admissions (125, 38.3%) were decompensations. One in three (114) had been booked in advance; none of these was a decompensation, yet they used 29.1% of all bed-days. Among 124 returns within 30 days, planned admissions outnumbered decompensations, 48 to 43. Counting all admissions tracked a patient’s decompensations only moderately (correlation 0.50) whereas counting urgent admissions tracked them closely (0.93). In crude analysis the sicker patients had fewer admissions, not more, partly because they were observed for a shorter time and partly because planned care goes to those well enough to receive it. A liver severity score, computable for 165 admissions, did not separate planned from urgent admissions or decompensations from other stays, even within one patient. Readmission was not the same thing as decompensation.

## Introduction

Alcohol-related cirrhosis is a major cause of liver-related death and generates substantial inpatient use across gastroenterology and internal medicine.[1–4] Hospital readmission is therefore widely used as a marker of disease burden, a quality-of-care outcome, and a target for intervention.[5–11] Yet an admission is an encounter, not a disease phase: the same patient may be admitted for new ascites, for planned endoscopy, for alcohol withdrawal, or for something unrelated. In alcohol-related disease the mixture is likely to be widest, because admissions for withdrawal or intoxication add a category that is neither decompensation nor planned care, and because abstinence can recompensate the liver, so that a later admission need not mean the disease has advanced. Readmission studies use a mixture of clinical and administrative data.[5–8] These studies are essential for estimating population rates, but routinely collected data cannot always determine whether a repeated hospitalization represents acute hepatic decompensation, and diagnostic codes for cirrhosis and its complications are themselves imperfect.[12,13] This distinction matters because the clinical course of cirrhosis is longitudinal,[14] whereas a hospital record describes a single encounter. If planned and non-decompensating admissions accumulate over time, hospitalization count may progressively diverge from the burden of acute decompensation in the same patient.

The same problem affects short-term readmission metrics. A return within 30 days is often read as clinical deterioration or as a failure of transitional care, even though some returns are scheduled. Claims-based quality measures therefore use algorithms to identify planned readmissions,[15,16] and chart-reviewed work in cirrhosis has shown that many 30-day readmissions are not preventable.[17] Frequent readmission has also been linked to identifiable patient-level risk factors.[18] Two questions remain open. The first is how far an admission count actually tracks the burden of decompensation, and whether it does so in the direction usually assumed, since planned care may accumulate in patients well enough to receive it. The second is whether a severity score such as the Model for End-Stage Liver Disease (MELD) could stand in for a record review by telling an urgent, decompensating admission from a scheduled one.

We therefore studied a deliberately selected cohort of patients with alcohol-related cirrhosis who had been hospitalized more than once. We did not aim to estimate population readmission risk. We asked what repeated hospitalizations consist of; how closely total and urgent admission counts reflect definite decompensation burden within patients; how those counts relate to severity at the index admission; whether an episode-level MELD distinguishes the kinds of admission that occur; and what an all-cause 30-day return represents. Whether the probability of decompensation changed across successive admissions was explored secondarily, and comparisons with routine administrative and coded data were secondary.

## Materials and methods

### Design and cohort

This retrospective cohort study was conducted at the Department of Gastroenterology, Hepatology, Toxicology and Internal Diseases of the University Hospital in Krakow, Poland. Departmental records were screened for adults with alcohol-related cirrhosis whose index admission fell between January 2013 and January 2026 and who had been hospitalized more than once. Repeated hospitalization was therefore an entry criterion rather than an outcome. The cohort cannot estimate the probability of first readmission, predictors of becoming a patient with repeated hospitalizations, or population readmission rates.

Of 73 patients assessed, 4 were excluded because hepatic failure was documented without established cirrhosis. Of the remaining 69, 7 were excluded because the hepatic phenotype was competing or uncertain, and 1 patient died during the index hospitalization and therefore had no recurrent admission. The primary cohort comprised 61 patients with established alcohol-related cirrhosis and 326 hospitalizations occurring after the first captured, index admission (Fig 1). The series spans thirteen years, so the composition reported here is not a snapshot of one configuration of the service. For every included patient, all consecutive admissions to the department until the end of follow-up were included; no episode was left out because of its reason, its length, or how clear the record was, and episodes that could not be classified were kept as indeterminate rather than dropped. Follow-up began at discharge from the index hospitalization and was administratively censored on July 19, 2026. Hospitalizations outside the study system were unavailable.

**Fig 1.**
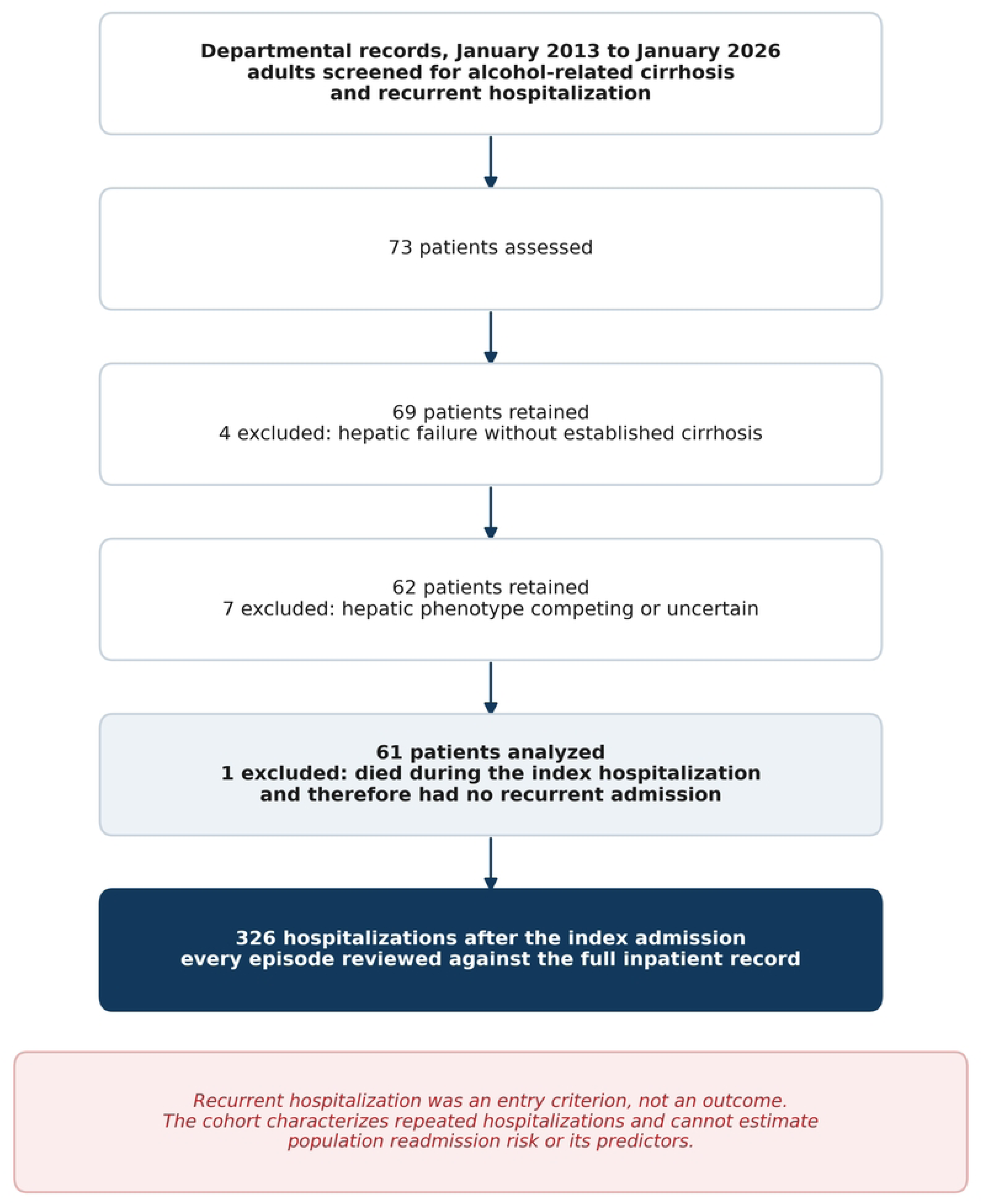
Cohort assembly and analytic framework. Patients entered the study because they had already been hospitalized more than once; the study therefore characterizes repeated hospitalizations rather than estimating population readmission risk. The 61-patient primary cohort contributed 326 post-index hospitalizations.

### Clinical review of recurrent hospitalizations

Each of the 326 post-index hospitalizations underwent clinician review of the full de-identified record. One physician reviewed all episodes; a second independently reviewed a random sample of 62 to assess reproducibility, and ratings in that sample were not reconciled. Both were blinded to the structured decompensation flags in the departmental database. The main analysis uses the resulting classification of 125 definite decompensations, 128 episodes without decompensation, and 73 indeterminate episodes; indeterminate episodes were retained as a distinct category and never forced into a binary label.

Acute hepatic decompensation was classified using a study operational definition, applied uniformly to all episodes and informed by European Association for the Study of the Liver guidance and the CANONIC framework.[19,20] Definite decompensation required new or worsening ascites, hepatic encephalopathy, gastrointestinal or variceal hemorrhage, spontaneous bacterial peritonitis, hepatorenal syndrome or acute kidney injury, or rising jaundice to be documented as a reason for admission. Stable chronic findings, scheduled procedures such as elective variceal band ligation performed for secondary prophylaxis,[21] and alcohol withdrawal or intoxication without hepatic decompensation did not qualify; precipitating events were recorded descriptively rather than used to define decompensation.[22] Admission mode was classified separately as urgent, planned, or indeterminate according to whether the hospitalization was clinically unscheduled, occurred on an agreed date, or could not be reconstructed from the record. The nature of any variceal event and a single dominant reason for admission were also recorded.

Length of stay was calculated from admission and discharge dates because the departmental structured length-of-stay field was empty. Separately, where a corresponding hospital administrative record was available, its admission-mode field was retained as an administrative variable and was not used to overwrite the clinician-reviewed mode. Structured flags for ascites, encephalopathy, varices, spontaneous bacterial peritonitis, hepatorenal syndrome or acute kidney injury, and jaundice were used only in secondary analyses. Operational definitions and source hierarchy are provided in Supplementary material, Methods S1.

Model for End-stage Liver Disease (MELD) scores were recomputed from laboratory values rather than transcribed. A patient-level score was calculated for the index admission, and an episode-level score for every post-index hospitalization with a complete contemporaneous bilirubin, international normalized ratio and creatinine panel; scores were not imputed and values from different days were not combined. No dialysis correction was applied because dialysis timing was unavailable in the laboratory source. A score was computable for 165 of the 326 hospitalizations and for 47 of the 61 patients at the index admission. At patient level, each admission count was correlated with index MELD (Spearman, patient-resampled 95% confidence intervals); as a sensitivity analysis for unequal observation time, counts were divided by person-years after index discharge and the correlation recalculated on the rates. Episode-level scores were compared between admission types by the difference in medians with patient-resampled intervals, and within patients with a score on both types by the median within-patient difference (each patient contributing the median of each type), the Hodges-Lehmann shift with its 95% confidence interval, and the Wilcoxon signed-rank test.

### Statistical analysis

Analyses were descriptive and longitudinal. The unit of analysis for the classification results is the hospitalization (n = 326) and for the utilization results the patient (n = 61). Continuous variables are reported as median (Q1-Q3), where Q1 and Q3 denote the first and third quartiles, and categorical variables as n (%). Because repeated hospitalizations are clustered within patients, confidence intervals for episode-level proportions and the secondary comparisons with routine data were obtained by patient-level cluster bootstrap with 20 000 resamples and a fixed random seed of 20260819.

An exploratory longitudinal analysis asked whether the probability that a hospitalization represented definite decompensation changed with successive admissions within the same patient. Indeterminate episodes were excluded, as were patients without within-patient variation in decompensation status. Admission sequence number and the decompensation indicator were centered within patient and their pooled correlation calculated; significance was assessed by 20 000 within-patient permutations of the decompensation labels.

A recurrent hospitalization was considered a 30-day return when it began 0 to 30 days after discharge from the preceding hospitalization, including the interval from the index discharge to the first recurrent admission. The clinical composition of all 30-day returns was described. For the exploratory comparison according to the mode of the preceding recurrent hospitalization, first recurrent admissions were excluded because the index hospitalization did not undergo the same post-index clinical review. The difference between return proportions after planned and urgent preceding admissions was summarized with a patient-cluster bootstrap confidence interval. At patient level, we summarized the fractions of repeated hospitalizations that were definite decompensations, urgent, or planned, and defined admission pattern from hospitalizations with a determined mode as urgent-only, planned-only, or mixed. The association between each patient’s number of repeated hospitalizations and number of definite decompensations was quantified by Spearman correlation and compared with the analogous correlation for urgent-admission count by paired patient bootstrap. Analyses were performed in Python 3.11 with NumPy 1.26, SciPy 1.11 and pandas 2.1. No prognostic model was fitted, because the study was designed to characterize repeated hospitalizations rather than to predict outcomes. There was no a priori sample size calculation; every patient meeting the entry criteria in the study window was included, and the resulting size was fixed by the cohort rather than chosen. The analyses are descriptive and are reported as estimates with confidence intervals rather than as a family of significance tests, and no adjustment was made for multiple comparisons; the individual P values should be read in that light, and the longitudinal and severity analyses are labeled exploratory where they are reported. Reporting follows STROBE and RECORD.[23,24]

### Ethics statement

The study was approved by the Research Ethics Committee of the Jagiellonian University Medical College (Komisja ds. Etyki Badan Naukowych Uniwersytetu Jagiellonskiego - Collegium Medicum), Krakow, Poland, by resolution 414/2026 adopted on July 2, 2026 (application 1072.6120.2.299.2026). All methods were carried out in accordance with the Declaration of Helsinki and with relevant institutional guidelines and regulations. The Research Ethics Committee waived the requirement for informed consent, as recorded in the approved study application, because the study was a retrospective analysis of existing de-identified hospital records and involved no contact with patients. Data were accessed for research purposes between 07/07/2026 and 20/07/2026. The authors who extracted the data had access to information that could identify individual patients during extraction. The records were de-identified before clinical review and analysis, and the study dataset contains no direct identifiers. The study is reported in accordance with the STROBE and RECORD statements; the completed STROBE checklist is provided in S1 File and the RECORD checklist in S2 File.

### Use of large language models

Large language model assistants (Chat GPT 5.6) were used to help with language editing. All resulting text and code were reviewed, edited and verified by the authors. The tools were not used for clinical record review, episode classification, statistical decisions, data interpretation, or the study conclusions. The authors take full responsibility for the manuscript and the analyses.

## Results

### Cohort

The source database contained 427 hospitalizations among the 69 patients initially retained after the first screening step: 69 index admissions and 358 subsequent hospitalizations. After the phenotype and recurrent-use exclusions, 61 patients contributed the 326 post-index hospitalizations analyzed here. Median age at the index admission was 55 years (Q1-Q3, 48-63), 48 patients (79%) were men, and 14 (23%) died during observed follow-up. Median follow-up was 491 days (Q1-Q3, 303-855). Baseline characteristics are summarized in Table 1.

**Table 1.**
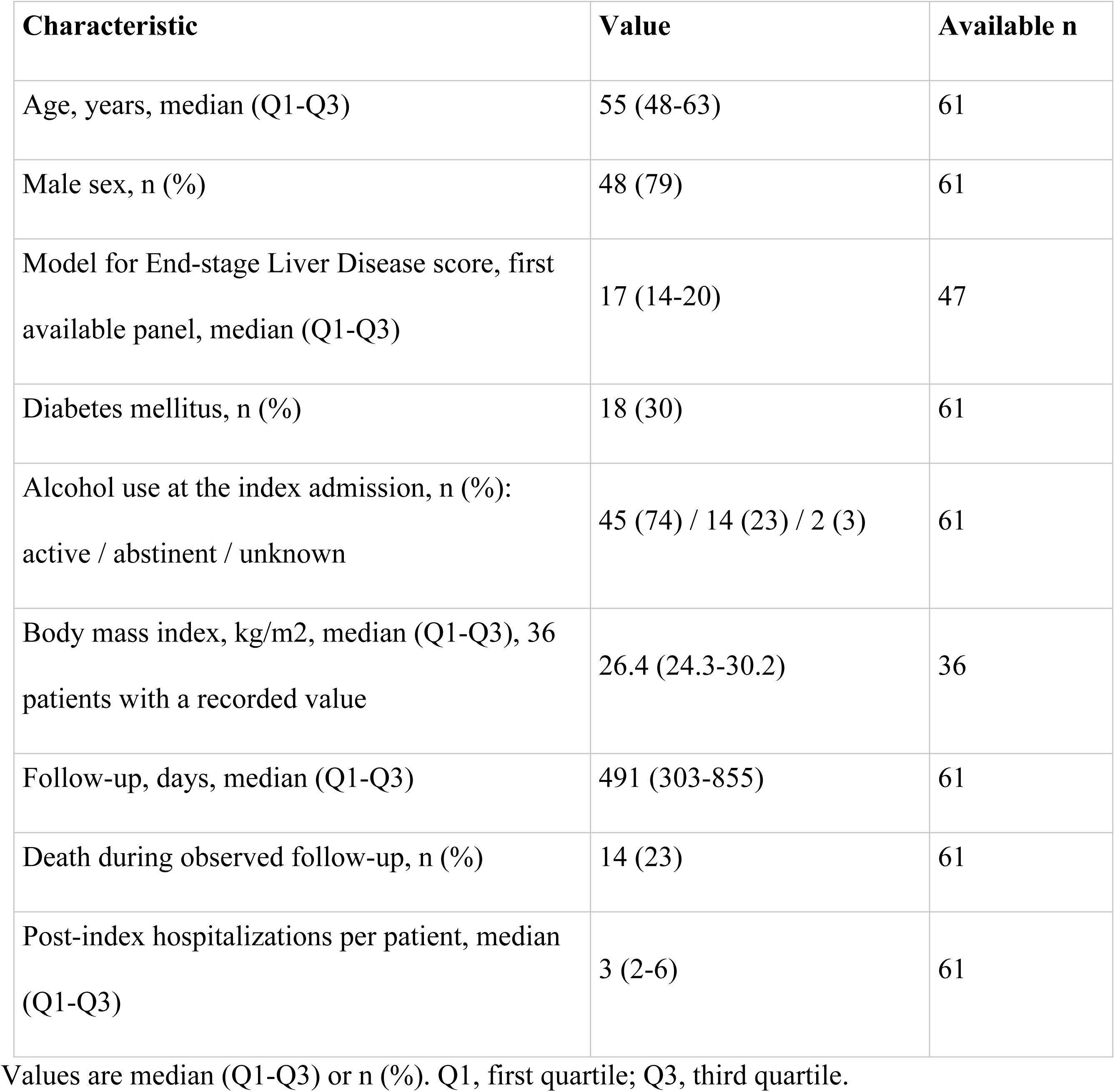
Characteristics of the primary cohort at the index admission (n = 61)

| Characteristic | Value | Available n |
| --- | --- | --- |
| Age, years, median (Q1-Q3) | 55 (48-63) | 61 |
| Male sex, n (%) | 48 (79) | 61 |
| Model for End-stage Liver Disease score, first available panel, median (Q1-Q3) | 17 (14-20) | 47 |
| Diabetes mellitus, n (%) | 18 (30) | 61 |
| Alcohol use at the index admission, n (%):<br>active / abstinent / unknown | 45 (74) / 14 (23) / 2 (3) | 61 |
| Body mass index, kg/m<sup>2</sup>, median (Q1-Q3), 36 patients with a recorded value | 26.4 (24.3-30.2) | 36 |
| Follow-up, days, median (Q1-Q3) | 491 (303-855) | 61 |
| Death during observed follow-up, n (%) | 14 (23) | 61 |
| Post-index hospitalizations per patient, median (Q1-Q3) | 3 (2-6) | 61 |

Values are median (Q1-Q3) or n (%). Q1, first quartile; Q3, third quartile.

### Clinical composition and inpatient burden

Of 326 recurrent hospitalizations, 125 (38.3%; patient-cluster bootstrap 95% confidence interval [CI], 28.4%-48.6%) were definite acute hepatic decompensations, 128 (39.3%; 95% CI, 28.5%-50.8%) were not decompensations, and 73 (22.4%; 95% CI, 16.5%-28.6%) remained indeterminate (Fig 2, Table 2). Admission mode was urgent in 143 episodes (43.9%), planned in 114 (35.0%), and indeterminate in 69 (21.2%). Under the definitions used, planned admission and definite acute decompensation did not overlap: all 114 planned admissions were classified as not representing acute decompensation. Because both classifications were assigned by the same reviewer from the same record, this separation is descriptive and is not an independent validation of admission mode. Of 143 urgent admissions, 124 (86.7%) were definite decompensations, while 68 of 69 admissions of indeterminate mode were also indeterminate for decompensation. Alcohol-related admissions without liver involvement accounted for 6 of the 326 hospitalizations.

**Fig 2.**
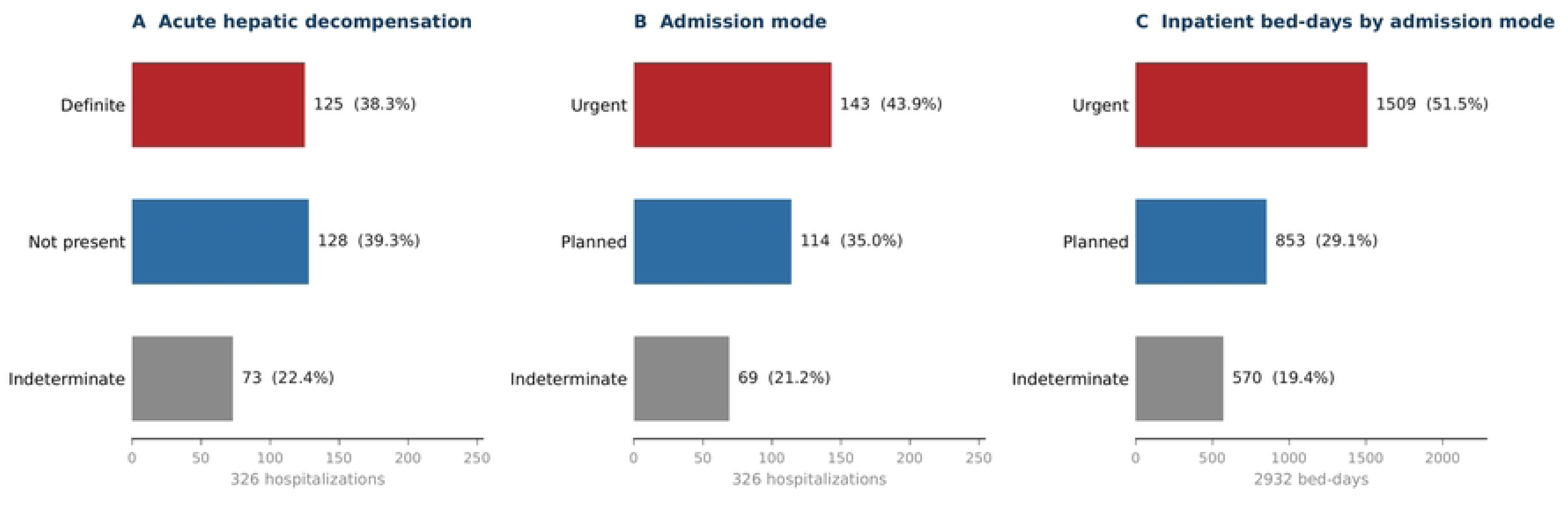
Clinical composition and inpatient burden of repeated hospitalizations. A, clinician-reviewed acute hepatic decompensation status for 326 hospitalizations. B, clinician-reviewed admission mode. C, distribution of 2932 recurrent inpatient bed-days by admission mode.

**Table 2.**
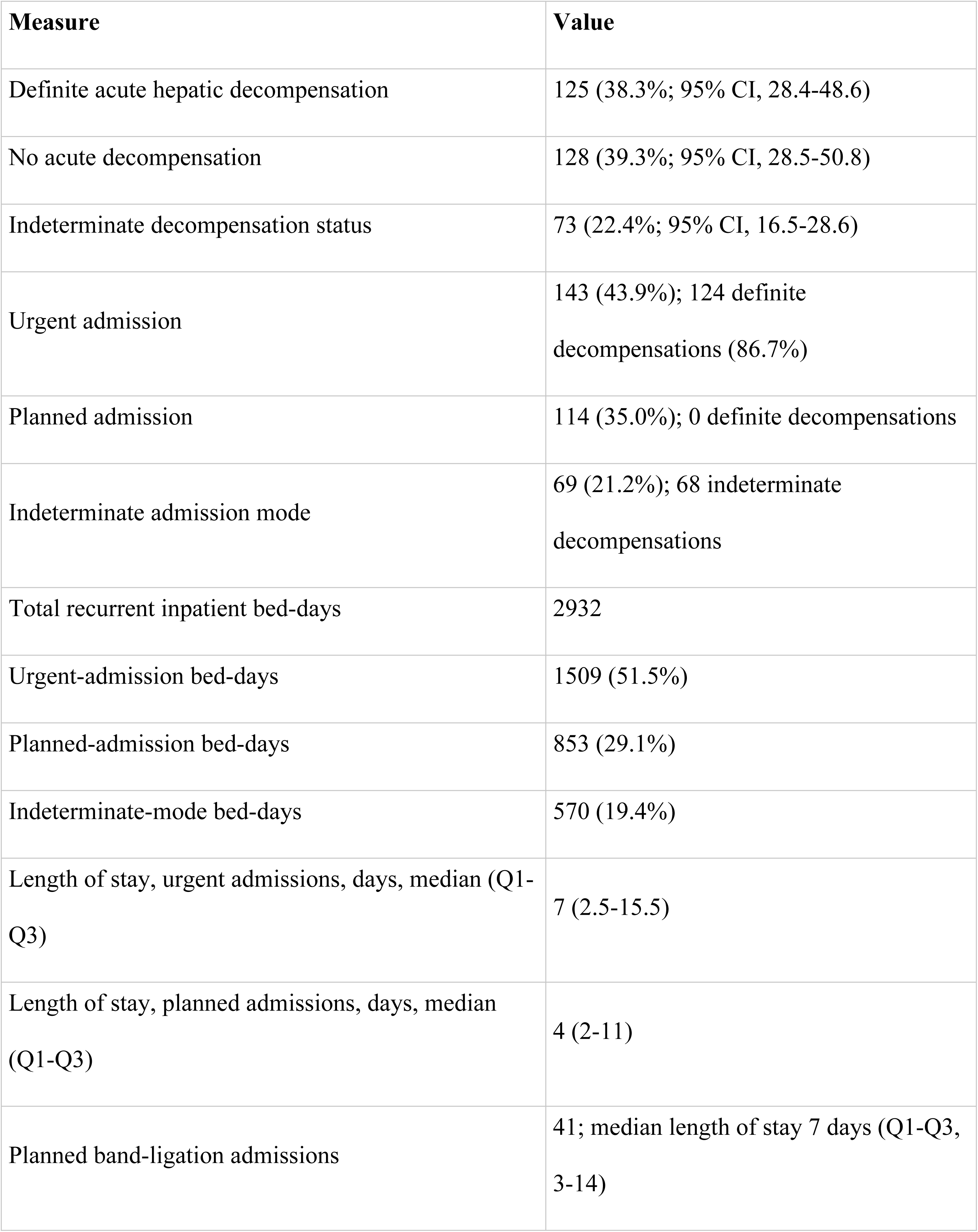
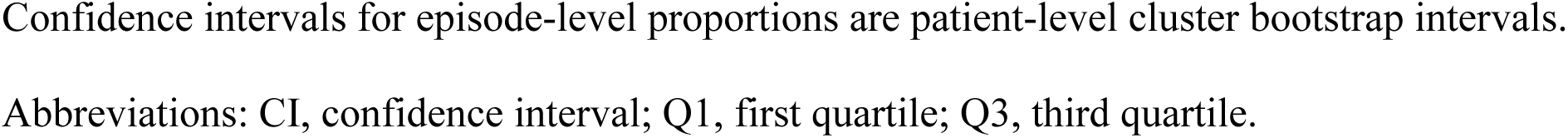
Clinical composition and inpatient burden of 326 recurrent hospitalizations.

| Measure | Value |
| --- | --- |
| Definite acute hepatic decompensation | 125 (38.3%; 95% CI, 28.4-48.6) |
| No acute decompensation | 128 (39.3%; 95% CI, 28.5-50.8) |
| Indeterminate decompensation status | 73 (22.4%; 95% CI, 16.5-28.6) |
| Urgent admission | 143 (43.9%); 124 definite decompensations (86.7%) |
| Planned admission | 114 (35.0%); 0 definite decompensations |
| Indeterminate admission mode | 69 (21.2%); 68 indeterminate decompensations |
| Total recurrent inpatient bed-days | 2932 |
| Urgent-admission bed-days | 1509 (51.5%) |
| Planned-admission bed-days | 853 (29.1%) |
| Indeterminate-mode bed-days | 570 (19.4%) |
| Length of stay, urgent admissions, days, median (Q1-Q3) | 7 (2.5-15.5) |
| Length of stay, planned admissions, days, median (Q1-Q3) | 4 (2-11) |
| Planned band-ligation admissions | 41; median length of stay 7 days (Q1-Q3, 3-14) |

Confidence intervals for episode-level proportions are patient-level cluster bootstrap intervals. Abbreviations: CI, confidence interval; Q1, first quartile; Q3, third quartile.

The 326 recurrent hospitalizations accounted for 2932 inpatient bed-days. Planned admissions accounted for 853 bed-days (29.1%), urgent admissions for 1509 (51.5%), and admissions with indeterminate mode for 570 (19.4%). Median length of stay was 7 days (Q1-Q3, 2.5-15.5) for urgent admissions and 4 days (Q1-Q3, 2-11) for planned admissions. Planned variceal band ligation was recorded in 41 hospitalizations and had a median length of stay of 7 days (Q1-Q3, 3-14), illustrating that planned care could still consume substantial inpatient resources in this center.

At patient level, 16 of 61 patients (26%) had no confirmed post-index decompensation despite recurrent hospitalization. Considering only admissions with a determined mode, 18 patients had urgent admissions only, 16 had planned admissions only, and 27 had both urgent and planned admissions. Thus, similar counts of recurrent hospitalization arose from clinically different utilization patterns.

### Within-patient trajectory

Among episodes with a definite decompensation classification, 28 patients showed within-patient variation and contributed 186 hospitalizations. After centering within patient, successive admission number correlated inversely with definite decompensation (r = -0.15; permutation P = 0.02) under the primary handling of indeterminate episodes (Supplementary Figure S1). The association was not robust to their alternative handling: counting them as non-decompensating gave r = -0.10 (34 patients, 256 episodes; P = 0.07) and counting them as decompensating gave r = -0.05 (36 patients, 259 episodes; P = 0.40); the three specifications are set out in Supplementary Table S3.

A definite post-index decompensation occurred in 45 of 61 patients (74%). The first occurred a median of 144 days after the index admission (Q1-Q3, 55-335), or 126 days after index discharge (Q1-Q3, 39-310). In 27 of those 45 patients the first recurrent hospitalization was already a definite decompensation; 18 had at least 1 recurrent hospitalization before their first confirmed decompensating episode. Across all 45 patients with a confirmed post-index decompensation, the median number of preceding recurrent hospitalizations was 0 (Q1-Q3, 0-1).

### What a 30-day return represented

There were 124 recurrent hospitalizations within 30 days of the preceding discharge, representing 38.0% of all 326 post-index hospitalizations. They were a mixed group: 43 (34.7%) were definite decompensations, 56 (45.2%) were not decompensations, and 25 (20.2%) were indeterminate. By admission mode, 52 (41.9%) were urgent, 48 (38.7%) planned, and 24 (19.4%) indeterminate (Fig 3, Table 3). Thus, almost two thirds of all 30-day returns were either not confirmed decompensations or could not be classified as such.

**Fig 3.**
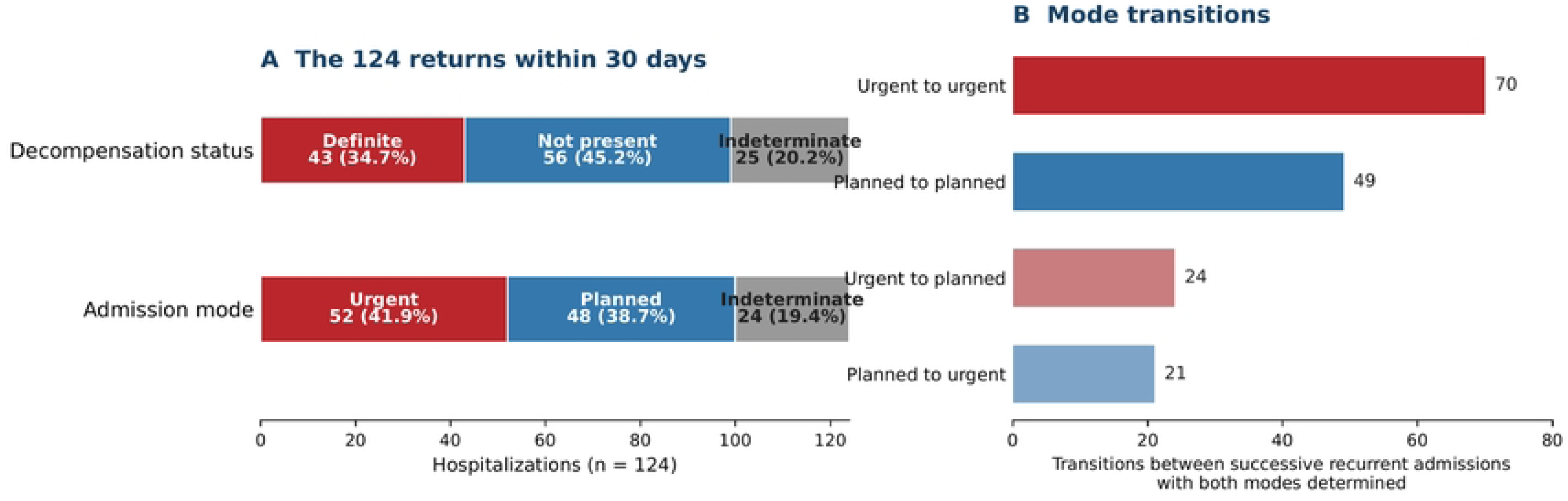
What a 30-day return hospitalization represented. A, decompensation status and admission mode among the 124 recurrent hospitalizations occurring within 30 days of the preceding discharge. B, transition counts between successive recurrent admissions for which both modes were determined; transitions involving the index admission are excluded.

**Table 3.** Longitudinal trajectory and 30-day return-hospitalization analyses.

| Measure | Value |
| --- | --- |
| Within-patient trajectory |  |
| Patients / episodes contributing within-patient contrast | 28 / 186 |
| Admission sequence vs definite decompensation | Within-patient $r = -0.15$; permutation $P = 0.02$ |
| Same analysis counting indeterminate episodes as non-decompensating | 34 patients / 256 episodes; $r = -0.10$; permutation $P = 0.07$ |
| Same analysis counting indeterminate episodes as decompensating | 36 patients / 259 episodes; $r = -0.05$; permutation $P = 0.40$ |
| Patients with $\geq 1$ post-index definite decompensation | 45 of 61 (74%) |
| Time to first post-index definite decompensation, days, median (Q1-Q3) | 144 (55-335) from index admission;<br>126 (39-310) from index discharge |
| Patients with $\geq 1$ recurrent hospitalization before first definite decompensation | 18 of 45 (40%) |
| Patients with no confirmed post-index decompensation | 16 of 61 (26%) |
| Admission-use phenotype based on determined-mode hospitalizations | Urgent-only 18; planned-only 16;<br>mixed 27 |
| 30-day return hospitalizations |  |
| All 30-day returns | 124 of 326 (38.0%) |
| Definite decompensation among 30-day returns | 43 of 124 (34.7%) |
| No decompensation among 30-day returns | 56 of 124 (45.2%) |
| Indeterminate decompensation among 30-day returns | 25 of 124 (20.2%) |
| Urgent / planned / indeterminate mode among 30-day returns | 52 / 48 / 24 |
| 30-day return after preceding planned recurrent admission | 41 of 89 (46.1%) |
| 30-day return after preceding urgent recurrent admission | 38 of 117 (32.5%) |
| Planned-minus-urgent difference | 13.6 percentage points (cluster-bootstrap 95% CI, -9.5 to 31.3) |

The planned-versus-urgent 30-day return comparison excludes first recurrent admissions because the preceding index hospitalization did not undergo the same post-index clinical review. The confidence interval crosses zero and the comparison is descriptive. Abbreviations: CI, confidence interval; Q1, first quartile; Q3, third quartile.

Among recurrent admissions of determined mode that were followed by another recurrent admission, a return within 30 days followed 41 of 89 planned admissions (46.1%) and 38 of 117 urgent admissions (32.5%). The difference was 13.6 percentage points, but the patient-cluster bootstrap 95% CI ranged from -9.5 to 31.3 percentage points; this comparison is therefore descriptive rather than confirmatory. Across recurrent-to-recurrent determined-mode pairs, urgent-to-urgent was the most frequent transition (70), followed by planned-to-planned (49), urgent-to-planned (24), and planned-to-urgent (21).

How admission counts, severity and routine data relate to decompensation burden At patient level, the number of all recurrent hospitalizations correlated moderately with the number of definite decompensations (Spearman rho = 0.50; bootstrap 95% CI, 0.28-0.69), whereas the number of urgent hospitalizations closely matched it (rho = 0.93; 95% CI, 0.84-0.98), as expected from the definitions, since urgent and decompensating are largely overlapping categories. The paired bootstrap difference was 0.42 (95% CI, 0.25-0.64).

Routine structured data said less. At least one departmental decompensation flag was present in 257 of 326 hospitalizations, of which 115 were definite decompensations, 93 were not, and 49 were indeterminate; 10 of the 125 definite decompensations carried no flag. Where both clinician-reviewed and hospital administrative admission mode could be compared, agreement was 61 of 108 episodes (56%). Detailed code-level performance, the administrative-mode comparison, and the independent duplicate review are presented in Supplementary material, Table S2 and Table S1. In the independent duplicate review of 62 episodes (19% of the series), the two clinicians agreed on 61.3% of decompensation assignments (Cohen kappa 0.45) and 54.8% of admission modes across all three categories; when both reached a definite answer, agreement was 93.8% for decompensation (30 of 32 episodes) and 90.0% for admission mode (27 of 30). Disagreement was therefore concentrated in the threshold for declaring a record insufficient rather than in the clinical judgment itself, which is why indeterminate episodes were kept as a category.

Admission count was inversely related to severity at the index admission. Among the 47 patients with a computable index MELD, the number of recurrent hospitalizations correlated negatively with that score (rho = -0.49; 95% CI, -0.69 to -0.25; Fig 4, Table 4). The association came from planned admissions (rho = -0.36; 95% CI, -0.63 to -0.05) and was absent for urgent admissions (rho = -0.26; 95% CI, -0.51 to 0.04) and for definite decompensations (rho = -0.17; 95% CI, -0.44 to 0.11). Across tertiles of index MELD (medians 12.5, 17.0 and 24.0), the median number of recurrent hospitalizations fell from 5.5 to 3.0 to 2.0 and the median number of planned admissions from 2.0 to 1.0 to 0.0, while the median number of definite decompensations remained between 1.0 and 1.5. Adjusting for observation time after index discharge removed the association for all hospitalizations (rho = -0.17; P = 0.25) but not for planned admissions (rho = - 0.29; P = 0.04), so the inverse relation of total count with severity is partly a matter of exposure, while the planned-admission component is not.

**Fig 4.**
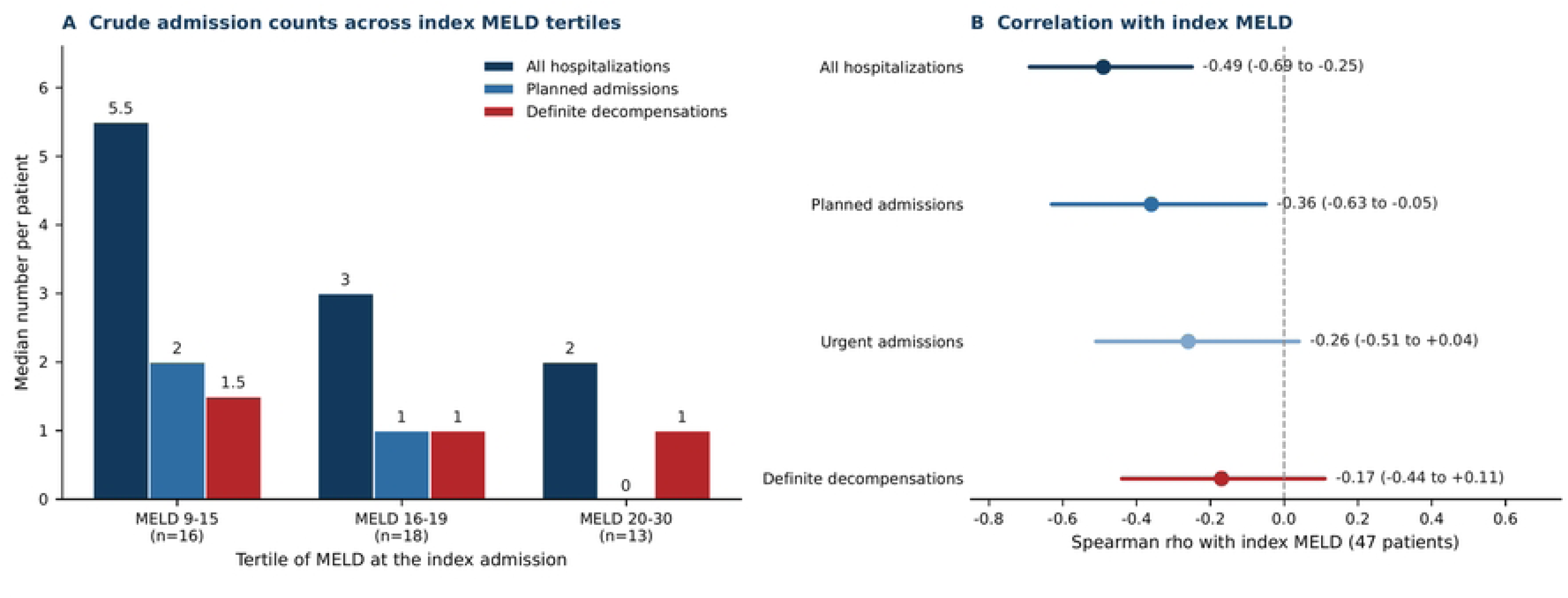
Crude admission counts and disease severity at the index admission. A, median number of all recurrent hospitalizations, planned admissions and definite decompensations per patient across tertiles of MELD at the index admission (47 patients with a computable score; tertile ranges 9 to 15, 16 to 19 and 20 to 30). B, Spearman correlation of each count with index MELD, with bootstrap 95% confidence intervals from 20 000 patient resamples. The crude inverse association of total admission count with index MELD weakened substantially after accounting for observation time; the planned-admission component remained inversely associated in that sensitivity analysis.

**Table 4.** Three tests of whether admission counts and a severity score capture what a hospitalization was for.

| Measure | Value |
| --- | --- |
| **Counts against decompensation burden (61 patients)** |  |
| All hospitalizations vs definite decompensations,<br>Spearman rho (95% CI) | 0.50 (0.28 to 0.69) |
| Urgent admissions vs definite decompensations,<br>Spearman rho (95% CI) | 0.93 (0.84 to 0.98) |
| **Association of counts with index MELD (47 patients)** |  |
| All hospitalizations, Spearman rho (95% CI) | -0.49 (-0.69 to -0.25) |
| Planned admissions, Spearman rho (95% CI) | -0.36 (-0.63 to -0.05) |
| Urgent admissions, Spearman rho (95% CI) | -0.26 (-0.51 to 0.04) |
| Definite decompensations, Spearman rho (95% CI) | -0.17 (-0.44 to 0.11) |
| Median hospitalizations by MELD tertile (low, middle,<br>high) | 5.5, 3.0, 2.0 |
| Median planned admissions by MELD tertile | 2.0, 1.0, 0.0 |
| Median definite decompensations by MELD tertile | 1.5, 1.0, 1.0 |
| **Episode-level MELD (165 hospitalizations)** |  |
| Urgent vs planned admissions, median (Q1-Q3) | 14 (11-18) vs 13 (10-18) |
| Difference in medians (95% CI, patient bootstrap) | 1.0 (-3.0 to 4.0) |
| Definite decompensation vs not, median (Q1-Q3) | 14 (11-18) vs 13 (10-18) |
| Difference in medians (95% CI, patient bootstrap) | 1.0 (-3.0 to 4.0) |
| Within-patient shift, urgent minus planned, 16 patients with both, Hodges-Lehmann (95% CI) | 0.50 (-2.75 to 2.00); Wilcoxon P = 0.49 |
| Within-patient shift, decompensation minus not, 14 patients with both, Hodges-Lehmann (95% CI) | 0.25 (-3.75 to 3.50); Wilcoxon P = 0.92 |
| Coverage of episode-level MELD, urgent vs planned admissions | 61% vs 42% |

Episode-level severity did not separate the kinds of admission that occurred. A MELD score was computable for 165 of the 326 hospitalizations (Table 4). The score did not differ between urgent and planned admissions (median 14 [11–18] versus 13 [10–18]; difference 1.0; 95% CI, -3.0 to 4.0) or between definite decompensations and hospitalizations classified as not decompensations (14 [11–18] versus 13 [10–18]; difference 1.0; 95% CI, -3.0 to 4.0), and the peak in-hospital score behaved identically. Within the same patient there was no evidence of a systematic difference either. Among the 16 patients with a score on both an urgent and a planned admission, the median within-patient difference (urgent minus planned, each patient contributing the median of that admission type) was 0.75 points, with a Hodges-Lehmann shift of 0.50 (95% CI, -2.75 to 2.00; Wilcoxon signed-rank P = 0.49); among the 14 patients with a score on both a decompensation and a non-decompensation the shift was 0.25 (95% CI, -3.75 to 3.50; P = 0.92).

## Discussion

In this cohort of 61 patients with alcohol-related cirrhosis, 326 consecutive hospitalizations divided into confirmed decompensation (38%), no decompensation (39%), and episodes the record did not allow us to classify (22%). Planned admissions made up a third of the total and none of them met the decompensation definition, yet they accounted for 29.1% of the inpatient days. Counting all hospitalizations tracked confirmed decompensation burden only moderately, while counting urgent admissions closely matched it, as expected from the definitions; separating urgent from planned care is therefore a practical way to split all-cause hospital use, not a validated substitute for reviewing the record. In crude analysis, admission count fell as severity at the index admission rose; the association came from planned admissions, weakened substantially once observation time was taken into account, and persisted for the planned component. And an episode-level severity score did not distinguish urgent from planned admissions or decompensations from other hospitalizations, even within the same patient. An all-cause admission count is therefore an imperfect measure of decompensation burden because it combines clinically different events, and a severity score does not correct it.

The crude inverse relation runs against the intuition that sicker patients are hospitalized more, and two mechanisms explain it. Part is time: patients with less advanced disease stay under observation longer and accumulate more admissions, which is why the association for total count weakened once observation time was taken into account. Part is care: elective procedural care, endoscopic surveillance and scheduled assessment accumulate in patients well enough to receive them, and this component survived adjustment. In this selected cohort, crude hospitalization count therefore depended partly on how long patients were followed and how much scheduled care they received, not only on acute liver deterioration.

Two features of this cohort are specific to alcohol-related disease. It was assembled on a joint gastroenterology, hepatology and toxicology ward, where admissions for withdrawal, intoxication and liver disease are seen by one team; even so, alcohol-related admissions without liver involvement were few (6 of 326), and what diluted the count was planned care, not alcohol. And in alcohol-related cirrhosis abstinence can recompensate the liver and drinking can reverse that, so the sequence of admissions in one patient need not track a steady decline.

That MELD did not separate the categories fits the argument rather than undermining it. MELD summarizes hepatic synthetic and renal dysfunction, a statement about disease stage; admission mode and acute decompensation are statements about the nature of the presenting event, and a patient admitted electively for band ligation may carry a higher score than one admitted urgently with new ascites. A severity score therefore cannot substitute for knowing why a patient was admitted.

Large cohorts have established readmission rates, predictors and reasons,[5–8] and our cohort, selected on repeated hospitalization, cannot estimate those probabilities. Its value lies in what a hospitalization represents rather than how often one occurs.

Of 124 returns within 30 days, 43 were definite decompensations and 48 were planned admissions. The obvious objection is that this is our problem: careful studies exclude planned admissions, and in many centers band ligation is a day case, so the 29% belongs to one service. Both are partly right; neither removes the finding. Studies with chart access can separate planned from urgent returns, as the recent REACT study did;[25] but most readmission figures for cirrhosis come from administrative data, where the separation is not visible, and the only tool built to recover it there found about half of the planned readmissions when tested against chart review (weighted sensitivity 49.8%, positive predictive value 58.7%).[15] And the planned share will differ between health systems. That is the point, not an objection: readmission rates are compared across cohorts and countries as if they counted the same event, and if the planned share ranges from a few percent to a third, they do not. Quality-indicator sets for cirrhosis and the programs built on them face the same ambiguity, because interventions at the transition out of hospital cannot touch admissions booked in advance,[26–28] and days at home has the same problem.[29] Before preventability is considered,[17] the event itself is heterogeneous: a dashboard that counts all-cause returns treats scheduled care as a failure of disease control.

Planned band-ligation admissions lasted a median of 7 days here, which is local organization. Recording whether an admission was scheduled and whether decompensation occurred would make later interpretation far more reliable.[29]

The within-patient trajectory analysis was exploratory: later admissions were modestly less likely to be definite decompensations under the primary specification (r = -0.15; permutation P = 0.02), but the association did not survive either alternative handling of indeterminate episodes.

The routine-data results show the same thing from another side. Structured decompensation flags were sensitive but nonspecific, and the administrative admission-mode field was not synonymous with clinical urgency. A code, an administrative pathway, a hospitalization count and clinician-reviewed acute deterioration are different variables. For utilization-based outcomes, the clinical interpretation therefore depends on which of these is meant.

This measurement problem is not confined to administrative data. In our recent systematic review of circulating adipokines in alcohol-related liver disease and MetALD, biomarker behavior varied across clinical phenotypes and disease stages, and the same principle holds at the biological level: a single proxy should not be taken as a complete picture of disease phase.[30]

### Study limitations

This study was designed as a complete measurement of one service rather than a sample of many, and its limits follow from that choice. Recurrent hospitalization was an entry criterion, so this is a selected high-utilization cohort rather than an inception cohort of cirrhosis, and the observed proportions are not population readmission rates. All patients came from a single ward and were classified throughout by one set of rules, which is what allows the composition to be read without the differences in coding practice, case definition and documentation standard that separate centers; the cost is that the proportions themselves are a property of this service, and local referral patterns and the use of inpatient beds for planned procedures will move them elsewhere. Hospitalizations outside the study system were unavailable, so the number of short-interval returns is a lower bound.

Clinical classification was retrospective and record-based. Seventy-three episodes remained indeterminate, and the duplicate review of 62 episodes showed moderate agreement driven mainly by different thresholds for declaring the record insufficient; we therefore retained indeterminate episodes rather than reclassifying them. The within-patient trajectory analysis included only the 28 patients providing within-person contrast and should not be extrapolated beyond them. The higher 30-day return proportion after planned than after urgent admissions had a wide confidence interval crossing zero and supports splitting the endpoint, not a causal claim. Finally, admission mode and decompensation status were derived from the same record by the same primary reviewer, so their strong association is descriptive and not an independent diagnostic validation.

A further limitation concerns the severity analyses. An index MELD was computable for 47 of 61 patients and an episode-level MELD for 165 of 326 hospitalizations, and the score was more often available for urgent admissions (61%) than planned ones (42%). The comparison between admission types is therefore conditioned on a sample enriched for urgent episodes, and the absence of a difference should be read with that in mind. No dialysis correction was applied because dialysis timing was unavailable. Alcohol use was recorded at the index admission only; drinking and abstinence over follow-up were not captured, so their contribution to the composition of later admissions could not be examined.

## Conclusions

Among patients with alcohol-related cirrhosis who had already been hospitalized repeatedly, neither counting admissions nor scoring severity captured what the hospitalizations were for. All-cause admission counts reflected confirmed decompensation burden only moderately and their relation to baseline severity depended on observation time, whereas planned admissions remained more frequent among patients with less advanced disease; counting urgent admissions tracked decompensation closely; and an episode-level severity score did not separate the kinds of admission that occurred. Studies and quality programs that use hospital utilization as an outcome should say plainly what they mean to measure and should separate planned from urgent admissions, because neither the count nor the score does it for them.

## Data Availability

The minimal data set underlying the findings, together with the analysis code that reproduces every reported value, is available in the Zenodo repository at https://doi.org/10.5281/zenodo.22035277 under controlled access. Public release of the individual-level data is restricted on ethical grounds. The dataset describes a small single-centre cohort of 61 patients. It is de-identified and contains no names, dates or locations, but the combination of indirect identifiers such as age, sex, sequence of admissions and clinical course could allow individual patients to be identified. Consent for open data sharing was not obtained, and the data remain under the custodianship of the study centre. The data are therefore held in Zenodo under controlled access. Requests for access may be sent to the Secretariat of the Chair and Clinic of Toxicology and Environmental Diseases, Jagiellonian University Medical College, Botaniczna 3, 31-503 Krakow, Poland, an institutional point of contact that does not depend on any individual author.

## Data availability

The de-identified patient-level and episode-level dataset, the independent duplicate ratings and the analysis code that support the findings of this study are available in the Zenodo repository, https://doi.org/10.5281/zenodo.22035277, under controlled access reviewed by the study center. Individual medical records and the free-text evidence recorded during clinical review are not publicly available because, in a cohort of this size, clinical narrative describing a specific admission could identify a patient; requests for access should be addressed to the Secretariat of the Chair and Clinic of Toxicology and Environmental Diseases, Jagiellonian University Medical College, Botaniczna 3, 31-503 Krakow, Poland, an institutional point of contact that does not depend on any individual author.

## Acknowledgments

None.

## Author contributions

K.M. conceived and designed the study, performed the data curation and formal analysis, and drafted the manuscript. A.Z. and D.Ł. contributed to the interpretation of the data. H.H. performed the clinical review of all 326 hospitalizations and contributed to data collection and interpretation. B.B.-S. performed the independent duplicate review of the reliability sample and contributed to the interpretation of the data. P.H. contributed to the design of the study and the interpretation of the data and provided resources and supervision. M.K. contributed to the concept and design of the study and the interpretation of the data and provided supervision. All authors revised the manuscript for important intellectual content, approved the final version, and agree to be accountable for all aspects of the work.

## Supporting information

S1 File. Supplementary information. Operational definitions and source hierarchy (Methods S1), reproducibility of the classification in the duplicate sample (Table S1), what the routine record held compared with the clinical review (Table S2), sensitivity of the within-patient trajectory analysis to indeterminate episodes (Table S3), the within-patient trajectory figure (Figure S1), and the completed STROBE checklist.

S2 File. RECORD checklist. Items of the RECORD extension for studies using routinely collected health data, with the location in the manuscript where each is reported.

